# Unravelling the influence of affective stimulation on functional neurological symptoms: A pilot experiment examining potential mechanisms

**DOI:** 10.1101/2023.08.23.23294462

**Authors:** Susannah Pick, L. S. Merritt Millman, Emily Ward, Eleanor Short, Biba Stanton, A.A.T.S. Reinders, Joel S. Winston, Timothy R. Nicholson, Mark J. Edwards, Laura H. Goldstein, Anthony S. David, Trudie Chalder, Matthew Hotopf, Mitul A. Mehta

## Abstract

**Background:** Differences in affective processing have previously been shown in functional neurological disorder (FND); however, the mechanistic relevance is uncertain. We tested the hypotheses that highly arousing affective stimulation would result in elevated subjective functional neurological symptoms (FNS), and this would be associated with elevated autonomic reactivity. The possible influence of cognitive detachment was also explored.

**Methods:** Individuals diagnosed with FND (motor symptoms/seizures; n=14) and healthy controls (HCs; n=14) viewed Positive, Negative, and Neutral images in blocks, whilst passively observing the stimuli (“Watch”) or detaching themselves (“Distance”). The FND group rated their primary FNS, and all participants rated subjective physical (arousal, pain, fatigue) and psychological states (positive/negative affect, dissociation), immediately after each block. Skin conductance (SC) and heartrate (HR) were monitored continuously.

**Results:** FNS ratings were higher after Negative compared to Positive and Neutral blocks in the FND group (p=0.002, η_p_^2^=0.386); however, this effect was diminished in the Distance condition relative to the Watch condition (p=0.018, η_p_^2^=0.267). SC and/or HR correlated with FNS ratings in the Negative-Watch and Neutral-Distance conditions (r-values: 0.527-0.672, p-values: 0.035-0.006). The groups did not differ in subjective affect or perceived arousal (p-values: 0.541-0.919, η_p_^2^: <0.001-0.015).

**Conclusions:** Emotionally significant events may exert an influence on FNS which is related to autonomic activation rather than altered subjective affect or perceived arousal. This influence may be modulated by cognitive detachment. Further work is needed to determine the relevance and neural bases of these processes in specific FND phenotypes.

---

**What is already known on this topic –** Functional neurological disorder (FND) samples show differences in affective responsivity and awareness; however, the direct influence of affective events on functional neurological symptoms (FNS) has not previously been demonstrated.

**What this study adds –** We piloted an experimental task allowing us to provide the first evidence of a direct influence of negative affective stimulation on momentary subjective FNS, which was associated with autonomic activation rather than changes in subjective affect or perceived arousal.

**How this study might affect research, practice or policy –** Our findings support models proposing roles for affective/autonomic mechanisms in FND, indicating that interventions aimed at improving awareness, integration and regulation of autonomic signals might be beneficial for some individuals with the disorder.

## Background

The pathophysiological mechanisms contributing to the generation of functional neurological symptoms (FNS) have not yet been explicated completely, although current models suggest possible interacting roles for altered attention, predictive processing, sense of agency, executive functioning, interoception and emotional processing[1]. Affective processing, specifically, has been a recurrent theme in theoretical models of functional neurological disorder (FND)[2–5].

The relevance of affective processing differences in FND has been broadly supported by empirical evidence. Many individuals with FND report emotionally salient experiences prior to the onset of the disorder or immediately preceding FNS occurrence/exacerbation[6–9]. Samples with FND exhibit altered autonomic and subjective (i.e., valence/arousal) responses to affective stimuli and differences in bodily/emotional awareness[4, 10–17].

Nevertheless, findings have been variable and significant methodological limitations identified[4, 18]. Notably, few studies have examined whether affective processing differences or autonomic arousal have a direct influence on sensorimotor function or FNS[19, 20].

Further research is needed to unravel possible interactions between affective processing, autonomic arousal, and FNS, with close attention to their temporal relationships. Examining these interactions will help determine whether affective processing differences and autonomic arousal exert a causal influence on FNS, rather than simply representing correlates of the disorder.

### Aims & hypotheses

We aimed to assess the feasibility and validity of an experimental task designed to examine the influence of affective stimulation and autonomic arousal on FNS severity.

We tested the following hypotheses[4]:

1. Individuals with FND would exhibit elevated autonomic arousal (skin conductance/heart-rate) versus healthy controls (HCs) during affective stimulation (positive/negative).
2. The FND group would report increased FNS severity immediately following affective stimulation, relative to a neutral control condition.
3. Autonomic arousal during affective stimulation would be associated with FNS severity ratings.
4. The relationship between autonomic arousal and subjective affect/perceived arousal would be weaker in the FND group than HCs.

There were also some exploratory aspects to this study. Dissociative tendencies are elevated in FND[21] and dissociation may contribute to the generation of FNS[17, 22]; therefore, we attempted to experimentally model dissociative states within the task. Pain and fatigue are common complaints in FND[23, 24] and may share common underlying mechanisms[25]. We included momentary probes to assess dissociation, pain and fatigue within the task, to examine the influence of affective stimulation on these other common symptoms.

## Methods

This experiment was part of a larger pilot study investigating aetiological factors and mechanisms in individuals with FND with motor symptoms and seizures. The study was approved by the King’s College London High Risk Research Ethics Committee (HR/DP-21/22-28714). Data were collected from July-October 2022.

### Participants

Fourteen participants diagnosed with FND with motor symptoms (n=11) or seizures (n=3) as their primary FNS were compared to HCs (n=14). This sample size was considered adequate to evaluate the feasibility of the paradigm and approximate effect sizes.

The recruitment/screening processes and eligibility criteria for the study are detailed in Supplementary Table 1 and previous publications[16, 26]. Participants with functional motor symptoms were asked to specify their primary motor symptom and those with functional seizures were asked to identify their most consistent premonitory symptom.

Individuals with functional seizures without premonitory symptoms were ineligible. All participants were reimbursed with a £50 shopping voucher.

### Materials & measures

#### Self-report measures

Validated questionnaires (Supplementary Table 2) assessed adverse life events, dissociative tendencies, anxiety, depression, alexithymia, and physical symptom burden. A bespoke Functional Neurological Symptoms Questionnaire (Supplementary Table 3) captured the range, severity and impact of FNS experienced in the FND sample.

#### Cognitive functioning

The two-subtest version of the Wechsler Abbreviated Scale of Intelligence–Second edition (WASI-II)[27] was administered to estimate full-scale intelligence-quotient (IQ).

#### Affective images task

The experiment had a mixed between- and within-groups design. The within-groups factors were image-type (Positive/Negative/Neutral) and task-instruction (Watch/Distance).

Participants were asked to either passively observe the images (Watch) or voluntarily detach themselves (Distance) (Supplementary Table 4). Affective images were selected from the International Affective Picture System[28] based on normative valence and arousal ratings (Supplementary Table 5).

The experiment was administered using E-Prime 3.0 (https://pstnet.com/products/e-prime/), consisting of 12 blocks of 10 images. We adopted a block-design to induce longer-term changes in emotional state and autonomic arousal than event-related designs[29]. Two blocks each of the following conditions were administered: Negative-Watch; Negative-Distance; Positive-Watch; Positive-Distance; Neutral-Watch; Neutral-Distance. The order of blocks was pseudorandomised.

Each block commenced with the task-instruction (Watch/Distance) presented for 2000ms. Ten images of the same type (Positive/Negative/Neutral) were then presented in a random order (6000ms each), all preceded by a fixation cross (500ms), and the word ‘Watch’ or ‘Distance’ (1000ms). Inter-stimulus intervals were jittered (1250-2000ms).

Participants completed momentary subjective assessments (Table 1) immediately after each block, followed by an inter-block interval (25-35s) during which the instruction ‘Rest’ appeared. Momentary FNS severity (FND group only), pain, fatigue and arousal were assessed with items developed by the research team and our FND Patient and Carer Advisory Panel. Items adapted from the Positive & Negative Affect Schedule[30] measured momentary affect, and items modified from the Clinician Administered Dissociative States Scale[31] assessed dissociative states. The order of the momentary probes was randomised, aside for the FNS ratings which always came first. Participants responded manually using a Likert-scale from 1 (Not at all) to 7 (Extremely).

**Table 1.** Subjective momentary assessments.

| **Dependent variable** | **Item wording: Right now, at the present moment....** |
| --- | --- |
| **Functional motor symptoms** | I am experiencing my primary FND symptom. |
| **Functional seizures** | I am experiencing my primary seizure warning symptom. |
| **Dissociation – depersonalisation** | • I feel disconnected from my own body.<br>• I feel separated from what is happening to me, like an actor in a movie, or a robot. |
| **Dissociation – derealisation** | • Things seem unreal to me, as if I am in a dream.<br>• It seems like I am looking at the world through a fog. |
| **Dissociation – amnesia** | • I cannot account for things that have recently happened.<br>• I feel spaced out, and/or have lost track of what is going on. |
| **Affect – positive** | I feel...<br>• Enthusiastic<br>• Determined<br>• Excited<br>• Alert<br>• Proud<br>• Strong |
| **Affect – negative** | I feel...<br>• Scared<br>• Upset<br>• Nervous<br>• Ashamed<br>• Irritable<br>• Hostile |
| **Arousal** | I feel bodily arousal.* |
| **Pain** | I am in bodily pain. |
| **Fatigue** | I feel tired. |

\* Instructions were given to participants to ensure that they understood the meaning of bodily arousal (i.e. sympathetic nervous system activation), with examples (e.g. dry mouth, racing heart, sweat response).

#### Psychophysiological measures

Psychophysiological measures were recorded using a Powerlab data acquisition system, with LabChart 8 software (https://www.adinstruments.com/). Recordings were acquired throughout the baseline period and experimental task, sampled at 1KHz.

#### Skin conductance

Skin conductance (SC) was measured with 8mm Ag/AgCl electrodes, filled with electrode paste and applied to the distal phalanges of the index and middle digits of the non-dominant hand[32]. SC was calibrated to measure a range of 0-50 microSiemens for each participant prior to the baseline recording.

##### Heart-rate

After skin preparation, electrocardiography electrodes were placed in an Einthoven triangle (LA/RA/LL). A range of 1-2 millivolts was adopted and adjusted to individual participants if necessary. Heart-rate (HR) in beats per minute (BPM) was computed from inter-beat intervals.

#### Procedure

Following a screening interview and online questionnaire pack described in detail elsewhere[16, 26], participants attended a laboratory testing session. All participants completed this experiment between 2-4pm in the same testing room, following approximately 1-2 hours of other cognitive/experimental tasks.

The psychophysiology electrodes were first attached and participants were seated for 3-5 minutes, before a 5-minute baseline recording. Participants then completed baseline subjective momentary assessments (Table 1) and were presented with written task instructions onscreen, followed by six practice images. The experimenter (SP) answered questions, checked participants’ understanding of the task, and remained present throughout the procedures.

#### Data processing and analysis

Data analyses were conducted in SPSS (v29, IBM) by SP and verified independently by LSMM. Values of 2.5 standard deviations above/below the group mean for each variable were considered outliers and winsorized. Hypothesis-driven tests were one-tailed (alpha p≤.05) and exploratory tests were two-tailed (alpha p≤.01). Effect sizes were Hedge’s g, r, or partial-eta squared.

Sociodemographic and clinical variables were analysed with between-group tests, including t-tests, Mann-Whitney, or chi-squared tests, as appropriate.

Momentary assessment scores were averaged across the two blocks for each condition. A two-way repeated-measures Analysis of Variance (ANOVA) assessed the influence of image-type (Positive/Negative/Neutral) and task-instruction (Watch/Distance) on subjective FNS ratings (FND group only). Three-way mixed ANOVAs were conducted for all other momentary subjective variables, with group as the between-group factor (FND/HC), and task-instruction (Watch/Distance) and image-type (Positive/Negative/Neutral) as within-group variables. Post-hoc t-tests adopted Bonferroni corrections.

Skin conductance (SC) and HR data were screened visually for artefacts and segments of contaminated data were excluded prior to analysis. SC and HR data for one participant from each group were excluded due to inadequate data quality. Baseline SC/HR scores were calculated from the mean values obtained during the last two-minutes of the baseline recording. Mean SC/HR scores were calculated for each block by subtracting baseline means from block means. Scores were averaged across the two blocks for each condition, analysed with three-way mixed ANOVAs (described above).

To examine the hypothesised relationship between momentary FNS severity and elevated autonomic arousal, correlations were computed between momentary FNS ratings and SC/HR for the conditions in which the highest FNS ratings were observed. Correlations were also carried out to test the hypothesis that the relationship between SC/HR and momentary subjective affect and arousal would be diminished in the FNS group relative to HCs.

Exploratory correlational analyses assessed possible relationships between key experimental dependent variables (momentary FNS ratings, SC/HR) and sociodemographic/clinical variables that differed significantly between groups. Pearson’s or Spearman’s coefficients were computed as appropriate.

## Results

### Sample characteristics (Table 2)

All participants in the FND group reported experiencing functional motor symptoms or functional seizures as their primary symptom, but they also reported at least one additional FNS.

**Table 2.** Demographic and clinical characteristics.

|  | **FND (n=14)** | **HC (n=14)** | **Test statistics (df)** | **p-value** | **Effect size** |
| --- | --- | --- | --- | --- | --- |
| **Age: M (SD)** | 39.2 (9.3) | 39.8 (11.1) | t(26)=-0.148 | 0.883 | g=0.054 |
| **Gender (female): n (%)** | 10 (71) | 11 (79) |  | 1.000* |  |
| **Handedness (right): n (%)** | 12 (86) | 13 (93) |  | 1.000* |  |
| **Body-mass index** | 28.0 (8.1) | 26.3 (5.5) | t(26)=0.646 | 0.524 | g=0.237 |
| **Relationship status (married/cohabiting): n (%)** | 9 (64) | 6 (43) |  | 0.450* |  |
| **Education (post-compulsory): n (%)** | 13 (93) | 14 (100) |  | 1.000* |  |
| **Ethnicity (white): n (%)** | 12 (86) | 9 (64) |  | 0.390* |  |
| **Employment status (employed/full-time education): n (%)** | 4 (29) | 13 (93) |  | 0.001* |  |
| **Full-scale IQ score: M (SD)** | 102.1 (14.6) | 105.6 (9.6) | t(26)=-0.764 | 0.452 | g=0.280 |
| **Primary FNS: n (%)** | FMS=11 (79)<br>FS=3 (21) |  |  |  |  |
| **Additional FNS:# n (%)** | Sensory=14 (100)<br>Dizziness=12 (86)<br>Speech/swallowing=9 (65)<br>Cognitive=11 (79) |  |  |  |  |
| **FNSQ Severity (1-7): M (SD)** | 3.9 (1.0) |  |  |  |  |
| **Impact (1-7): M (SD)** | 3.9 (0.8) |  |  |  |  |
| **Current mental health diagnosis: n (%)** | 8 (57) | 1 (7) |  | 0.013* |  |
| **Current physical health diagnosis (not FND): n (%)** | 10 (71) | 4 (29) |  | 0.057* |  |
| **Medication: n (%)** | 13 (93) | 4 (29) |  | 0.001* |  |

Notes: df=degrees of freedom; FMS=functional motor symptoms; FND=functional neurological disorder; FNS=functional neurological symptoms; FS=functional seizures; FNSQ= Functional Neurological Symptoms Questionnaire; IQ=intelligence quotient; M=mean; SD=standard deviation
\*Fisher's exact #All participants in the FNS group reported >1 FNS

The groups did not differ significantly on most possible confounding variables; however, a significantly greater proportion of the FND group reported mental health diagnoses and taking medication, and fewer of the FND group were in employment/full-time education. This FND sample reported significantly greater depression, anxiety, somatoform dissociation, depersonalisation, alexithymia, and physical symptom burden, compared to HCs (Supplementary Table 2).

### Subjective momentary assessments

#### Physical states (*Table 3*)

##### Functional neurological symptoms

The average momentary FNS severity rating at baseline was in the mild-moderate range. The task-based ANOVA yielded a significant main effect of image-type. FNS ratings were significantly higher following Negative compared to Positive (MD=0.55, p=0.009) and Neutral blocks (MD=0.38, p=0.048). However, FNS ratings did not differ for Positive and Neutral blocks (MD=0.18, p=0.599).

**Table 3.**
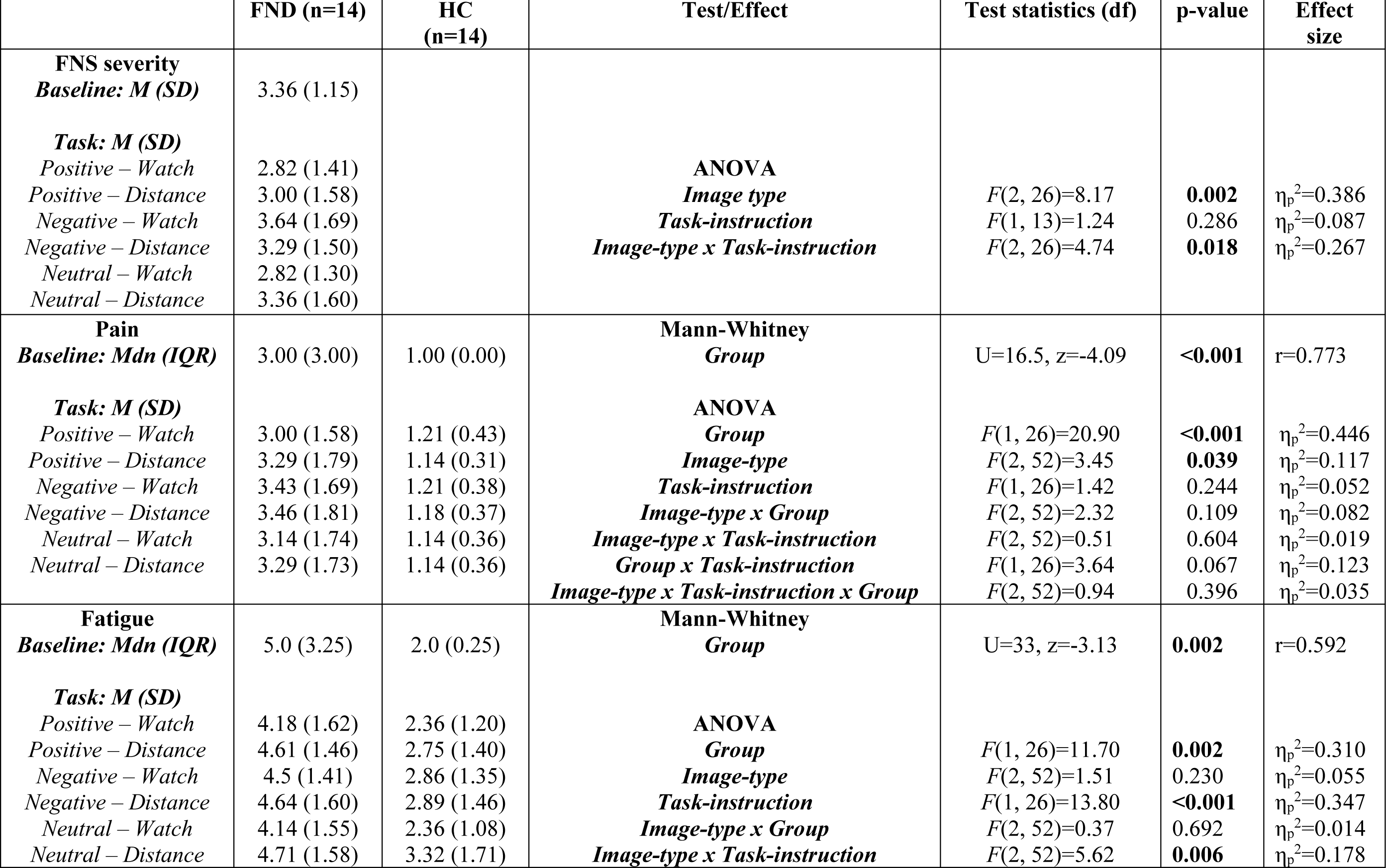
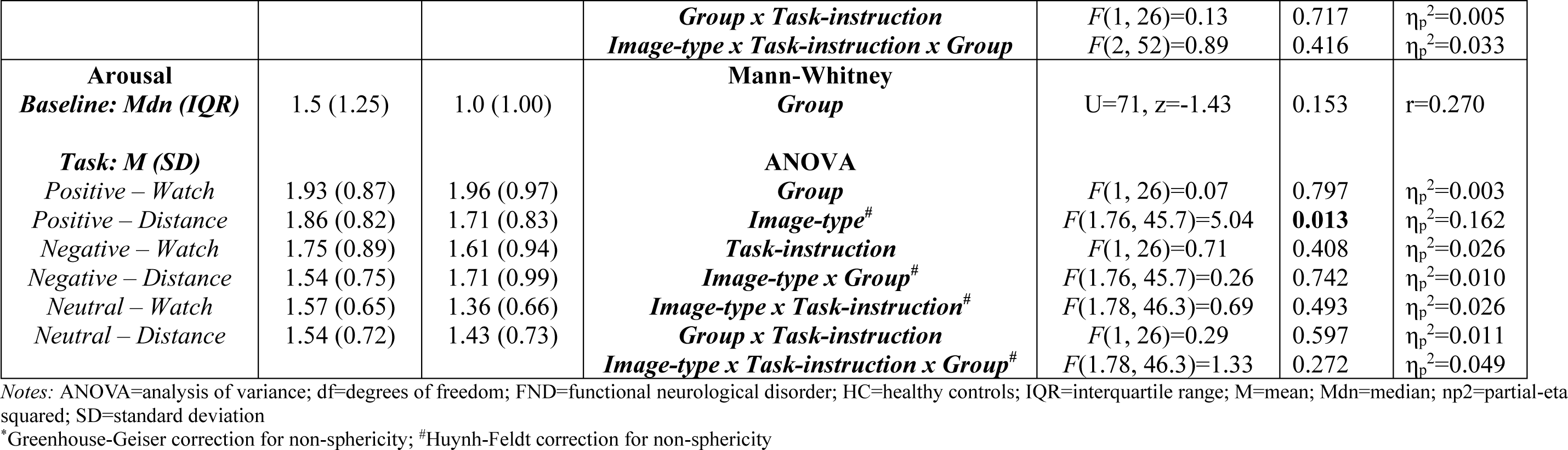
Subjective momentary assessment statistics – physical states.

|  | **FND (n=14)** | **HC (n=14)** | **Test/Effect** | **Test statistics (df)** | **p-value** | **Effect size** |
| --- | --- | --- | --- | --- | --- | --- |
| **FNS severity**<br>*Baseline: M (SD)*<br>*Task: M (SD)*<br>*Positive – Watch*<br>*Positive – Distance*<br>*Negative – Watch*<br>*Negative – Distance*<br>*Neutral – Watch*<br>*Neutral – Distance* | 3.36 (1.15)<br>2.82 (1.41)<br>3.00 (1.58)<br>3.64 (1.69)<br>3.29 (1.50)<br>2.82 (1.30)<br>3.36 (1.60) | | **ANOVA**<br>*Image type*<br>*Task-instruction*<br>*Image-type x Task-instruction* | *F*(2, 26)=8.17<br>*F*(1, 13)=1.24<br>*F*(2, 26)=4.74 | **0.002**<br>0.286<br>**0.018** | $\eta_p^2=0.386$<br>$\eta_p^2=0.087$<br>$\eta_p^2=0.267$ |
| **Pain**<br>*Baseline: Mdn (IQR)*<br>*Task: M (SD)*<br>*Positive – Watch*<br>*Positive – Distance*<br>*Negative – Watch*<br>*Negative – Distance*<br>*Neutral – Watch*<br>*Neutral – Distance* | 3.00 (3.00)<br>3.00 (1.58)<br>3.29 (1.79)<br>3.43 (1.69)<br>3.46 (1.81)<br>3.14 (1.74)<br>3.29 (1.73) | 1.00 (0.00)<br>1.21 (0.43)<br>1.14 (0.31)<br>1.21 (0.38)<br>1.18 (0.37)<br>1.14 (0.36)<br>1.14 (0.36) | **Mann-Whitney**<br>*Group*<br>**ANOVA**<br>*Group*<br>*Image-type*<br>*Task-instruction*<br>*Image-type x Group*<br>*Image-type x Task-instruction*<br>*Group x Task-instruction*<br>*Image-type x Task-instruction x Group* | U=16.5, z=-4.09<br>*F*(1, 26)=20.90<br>*F*(2, 52)=3.45<br>*F*(1, 26)=1.42<br>*F*(2, 52)=2.32<br>*F*(2, 52)=0.51<br>*F*(1, 26)=3.64<br>*F*(2, 52)=0.94 | **<0.001**<br>**<0.001**<br>**0.039**<br>0.244<br>0.109<br>0.604<br>0.067<br>0.396 | r=0.773<br>$\eta_p^2=0.446$<br>$\eta_p^2=0.117$<br>$\eta_p^2=0.052$<br>$\eta_p^2=0.082$<br>$\eta_p^2=0.019$<br>$\eta_p^2=0.123$<br>$\eta_p^2=0.035$ |
| **Fatigue**<br>*Baseline: Mdn (IQR)*<br>*Task: M (SD)*<br>*Positive – Watch*<br>*Positive – Distance*<br>*Negative – Watch*<br>*Negative – Distance*<br>*Neutral – Watch*<br>*Neutral – Distance* | 5.0 (3.25)<br>4.18 (1.62)<br>4.61 (1.46)<br>4.5 (1.41)<br>4.64 (1.60)<br>4.14 (1.55)<br>4.71 (1.58) | 2.0 (0.25)<br>2.36 (1.20)<br>2.75 (1.40)<br>2.86 (1.35)<br>2.89 (1.46)<br>2.36 (1.08)<br>3.32 (1.71) | **Mann-Whitney**<br>*Group*<br>**ANOVA**<br>*Group*<br>*Image-type*<br>*Task-instruction*<br>*Image-type x Group*<br>*Image-type x Task-instruction* | U=33, z=-3.13<br>*F*(1, 26)=11.70<br>*F*(2, 52)=1.51<br>*F*(1, 26)=13.80<br>*F*(2, 52)=0.37<br>*F*(2, 52)=5.62 | **0.002**<br>**0.002**<br>0.230<br>**<0.001**<br>0.692<br>**0.006** | r=0.592<br>$\eta_p^2=0.310$<br>$\eta_p^2=0.055$<br>$\eta_p^2=0.347$<br>$\eta_p^2=0.014$<br>$\eta_p^2=0.178$ |
| Affective stimulation, autonomic arousal and FNS |  |  |  |  |  |  |
| | | | *Group x Task-instruction*<br>*Image-type x Task-instruction x Group* | $F(1, 26)=0.13$<br>$F(2, 52)=0.89$ | 0.717<br>0.416 | $\eta_p^2=0.005$<br>$\eta_p^2=0.033$ |
| **Arousal**<br>**Baseline: Mdn (IQR)** | 1.5 (1.25) | 1.0 (1.00) | **Mann-Whitney**<br>**Group** | U=71, z=-1.43 | 0.153 | r=0.270 |
| **Task: M (SD)** |  |  | **ANOVA** |  |  |  |
| *Positive – Watch* | 1.93 (0.87) | 1.96 (0.97) | **Group** | $F(1, 26)=0.07$ | 0.797 | $\eta_p^2=0.003$ |
| *Positive – Distance* | 1.86 (0.82) | 1.71 (0.83) | **Image-type<sup>#</sup>** | $F(1.76, 45.7)=5.04$ | **0.013** | $\eta_p^2=0.162$ |
| *Negative – Watch* | 1.75 (0.89) | 1.61 (0.94) | **Task-instruction** | $F(1, 26)=0.71$ | 0.408 | $\eta_p^2=0.026$ |
| *Negative – Distance* | 1.54 (0.75) | 1.71 (0.99) | **Image-type x Group<sup>#</sup>** | $F(1.76, 45.7)=0.26$ | 0.742 | $\eta_p^2=0.010$ |
| *Neutral – Watch* | 1.57 (0.65) | 1.36 (0.66) | **Image-type x Task-instruction<sup>#</sup>** | $F(1.78, 46.3)=0.69$ | 0.493 | $\eta_p^2=0.026$ |
| *Neutral – Distance* | 1.54 (0.72) | 1.43 (0.73) | **Group x Task-instruction** | $F(1, 26)=0.29$ | 0.597 | $\eta_p^2=0.011$ |
| | | | **Image-type x Task-instruction x Group<sup>#</sup>** | $F(1.78, 46.3)=1.33$ | 0.272 | $\eta_p^2=0.049$ |

Notes: ANOVA=analysis of variance; df=degrees of freedom; FND=functional neurological disorder; HC=healthy controls; IQR=interquartile range; M=mean; Mdn=median; $\eta_p^2$ =partial-eta squared; SD=standard deviation
\*Greenhouse-Geiser correction for non-sphericity; <sup>#</sup>Huynh-Feldt correction for non-sphericity

There was a significant image-type x task-instruction interaction. FNS ratings were significantly elevated after Negative compared to both Positive (MD=0.82, p=0.035) and Neutral blocks (MD=0.82, p=0.032) in the Watch condition. In the Distance condition, FNS ratings were only higher for Negative compared to Positive (MD=0.29, p=0.017), but not Neutral blocks (MD=0.07, p=1.000). Mean ratings indicated that FNS were elevated after the Neutral-Distance blocks to a similar degree as the Negative-Distance blocks.

##### Pain

Pain ratings were significantly higher in the FND group compared to HCs at baseline and during the task. The task-based main effect of image-type was also significant. Pain ratings were higher following Negative (M=2.32, SE=0.236) compared to Positive (M=2.16, SE=0.228) or Neutral blocks (M=2.18, SE=0.232), although these differences were not significant following Bonferroni correction.

##### Fatigue

The FND group reported significantly greater fatigue than HCs at baseline and during the task. There was a significant main effect of task-instruction, revealing significantly higher fatigue ratings for the Distance condition than the Watch condition.

The interaction between image-type and task-instruction was also significant. Fatigue ratings were higher for Negative (M=3.67, SE=0.261) compared to Positive (M=3.27, SE=0.270) and Neutral blocks (M=3.25, SE=0.253) in the Watch condition. However, in the Distance condition, fatigue ratings were high across image-types and did not differ significantly (all>3.68).

##### Subjective arousal

The FND and HC groups reported comparable physiological arousal at baseline and during the task. There was a significant main effect of image-type on arousal ratings across the sample, with ratings significantly higher following Positive compared to Neutral blocks (MD=0.39, p=0.020), but not compared to Negative blocks (MD=0.21, p=0.440).

#### Psychological states (*Table 4*)

##### Affect

Positive affect ratings did not differ between groups at baseline or during the task. However, the main effects of image-type and task-instruction were significant. The main effect of image-type was due to elevated positive affect ratings following Positive compared to Negative (MD=0.37, p=0.003) and Neutral blocks (MD=0.24, p=0.035); positive affect ratings did not differ between Negative and Neutral blocks (MD=0.13, p=0.608). The main effect of task-instruction was due to significantly elevated positive affect ratings following the Watch condition compared to the Distance condition (MD=0.22, p<0.001).

**Table 4.**
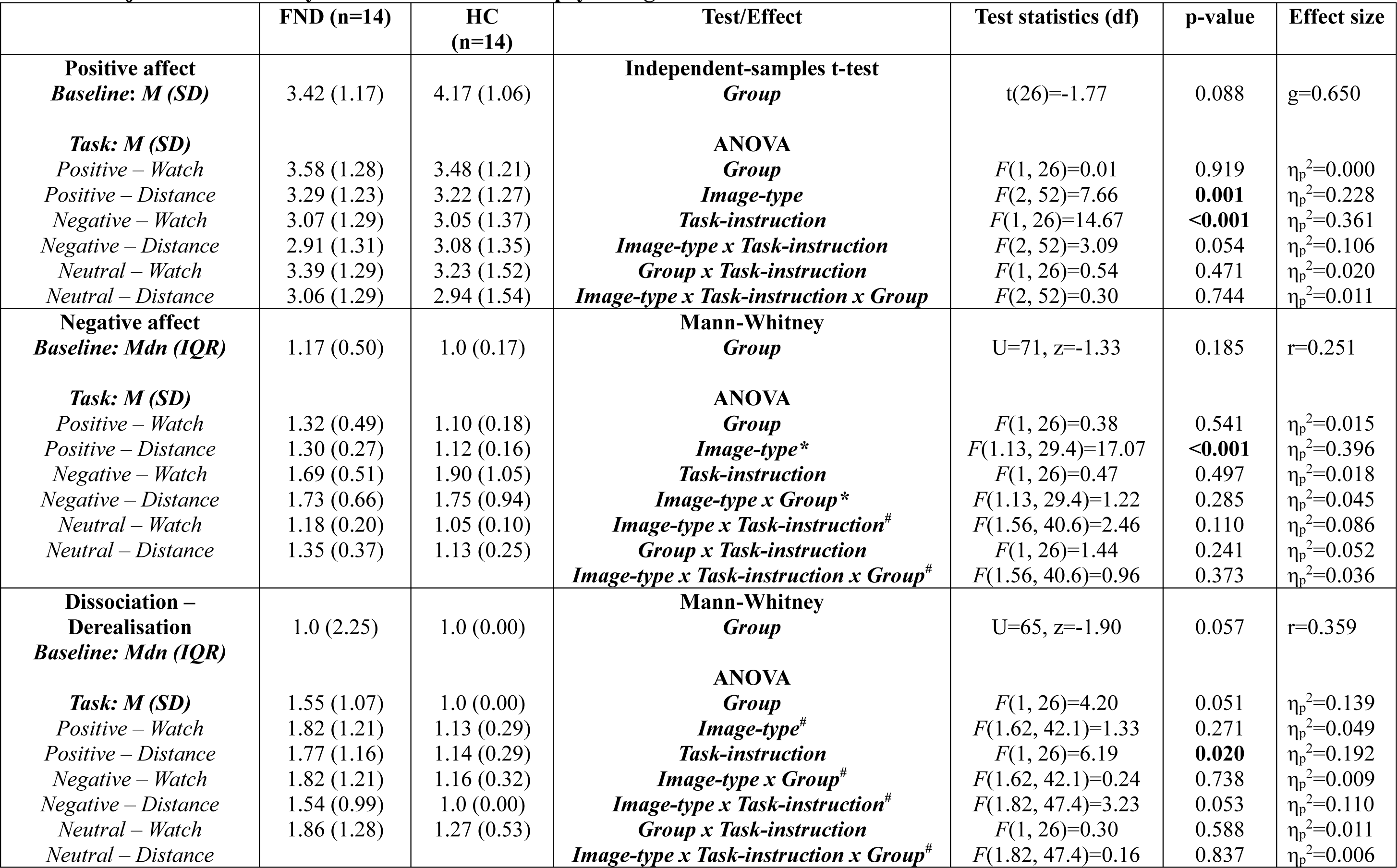
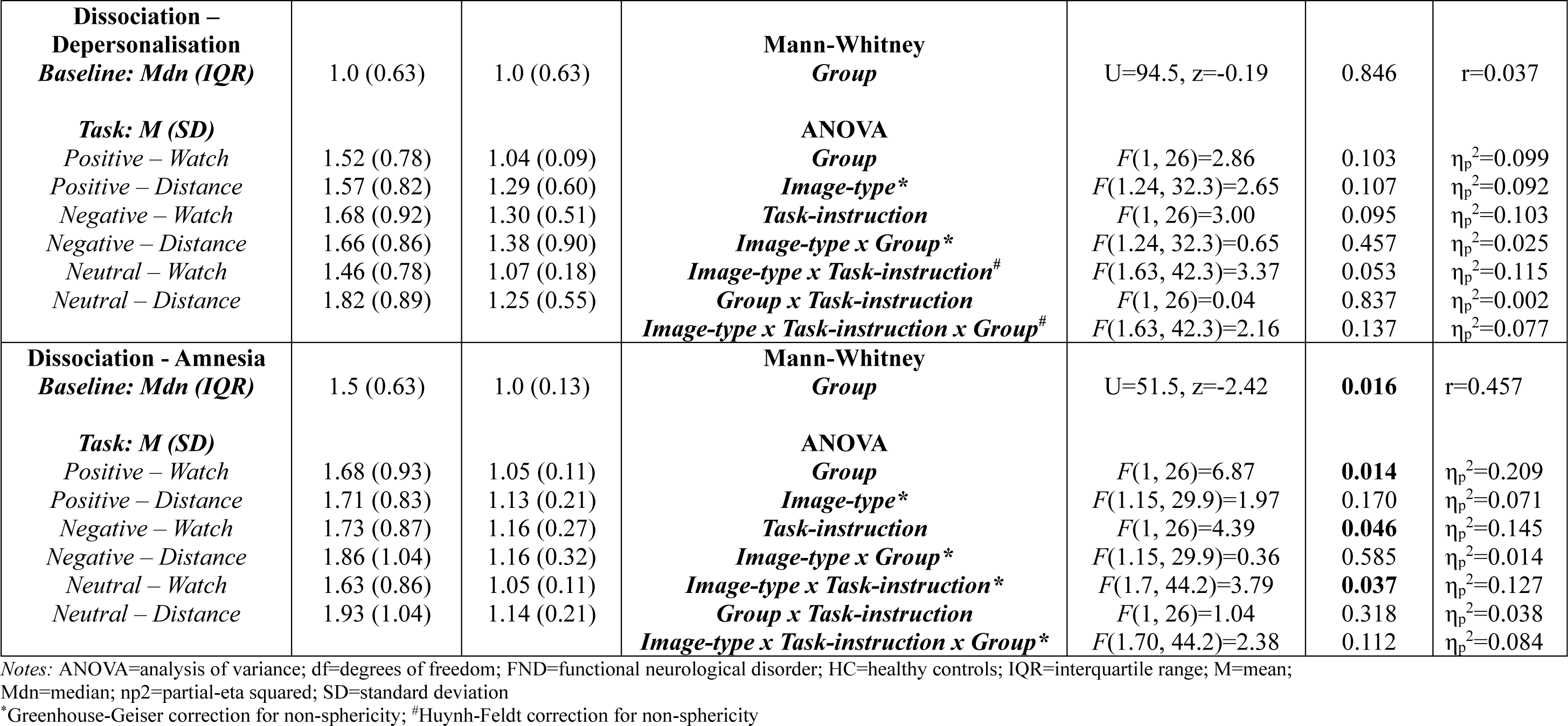
Subjective momentary assessment statistics – psychological states.

Negative affect did not vary between groups at baseline or in the task. The only significant task-based main effect was image-type, reflecting significantly higher negative affect ratings following Negative relative to Positive (MD=0.56, p<0.001) and Neutral blocks (MD=0.59, p<0.001). Ratings of negative affect did not differ between Positive and Neutral blocks (MD=0.03, p=1.000).

##### Dissociation

There were no significant group effects or interactions on depersonalisation ratings. There were also no group effects on derealisation ratings at baseline or in the task. However, the FND group reported significantly elevated amnesia compared to HCs at baseline and during the task.

There were significant main effects of task-instruction for derealisation and amnesia, with both elevated in the Distance compared to the Watch condition. There was a significant interaction between image-type and task-instruction on amnesia ratings. Post-hoc tests showed the effect of task-instruction was significant only for Neutral blocks, in which amnesia ratings were significantly higher in the Neutral-Distance condition compared to the Neutral-Watch condition (MD=0.20, p=0.016). There was no significant effect of task-instruction on amnesia ratings for Positive (MD=0.05, p=0.320) or Negative blocks (MD=0.06, p=0.193).

#### Psychophysiological measures (Table 5)

##### Skin conductance (SC)

There was no significant effect of group at baseline or during the task, and no interactions between group and other factors.

**Table 5.**
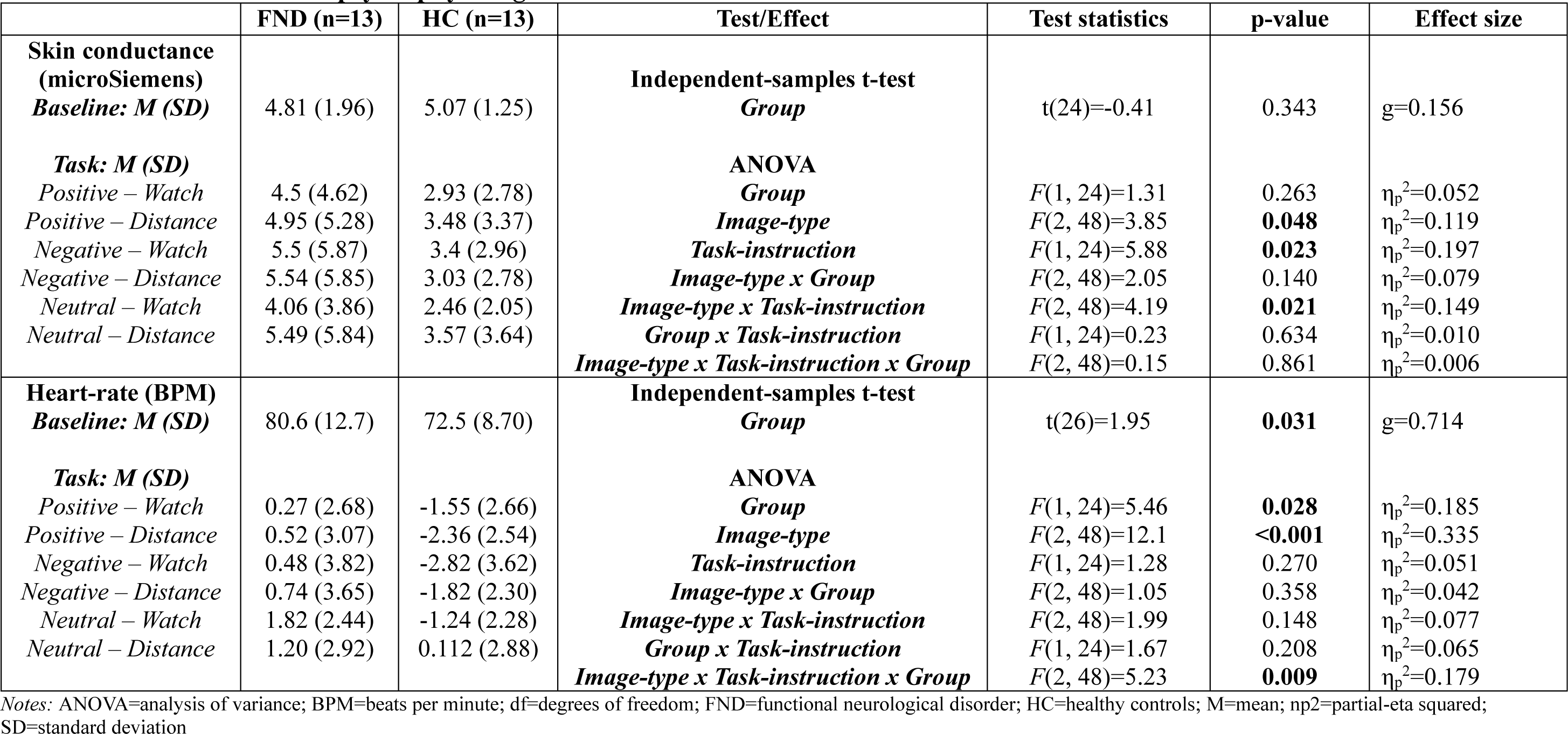
Statistical values for psychophysiological variables.

The main effect of task instruction was significant, reflecting significantly higher SC in the Distance condition, relative to Watch (MD=0.54, p=0.023). The main effect of image-type was also significant, with SC values highest for Negative blocks, followed by Positive and Neutral blocks.

There was a significant image-type x task-instruction interaction. The interaction was due to SC being significantly higher for Negative compared to Neutral blocks in the Watch condition (MD=1.19, p=0.013), but this difference was not significant in the Distance condition (MD=0.25, p=1.000).

A positive correlation was observed between SC and FNS ratings in the Negative-Watch (*r*=.628, p=.011) and Neutral-Distance conditions (*r*=0.517, p=0.035), but not the Negative-Distance condition (*r_s_*=0.253, p=0.202).

Positive and negative affect ratings were not correlated with SC in either group in the Negative-Watch, Negative-Distance and Neutral-Distance conditions. Arousal ratings were not correlated with SC in either group in the Negative-Watch and Negative-Distance conditions. However, in the Neutral-Distance condition, SC was correlated inversely with momentary arousal ratings in FND (*r_s_*=-0.606, p=0.014), but not HCs (*r_s_*=-0.167, p=0.293).

##### Heart-rate (HR)

Heart-rate was significantly elevated in the FND group compared to HCs during baseline and the task. Average HR accelerated during the task relative to baseline in the FND group, whereas it decelerated in HCs.

There was a significant main effect of image-type on HR during the task, with HR highest for Neutral compared to both Positive (MD=1.25, p<0.001) and Negative blocks (MD=1.33, p=0.004). There was also a significant group x task-instruction x image-type interaction. In the Positive condition, the FND group had higher HR than HCs for Distance (MD=2.87, p=0.016) but not Watch blocks (MD=1.82, p=0.095). In the Negative condition, the FND group exhibited higher HR for both Watch (MD=3.30, p=0.033) and Distance blocks (MD=2.57, p=0.043). For Neutral images, the FND group displayed elevated HR compared to HCs in the Watch blocks (MD=3.05, p=0.003) but not in the Distance blocks (MD=1.09, p=0.347).

In the FND group, HR was positively correlated with FNS ratings in the Negative-Watch (*r*=0.533, p=0.030) and Neutral-Distance conditions (*r*=0.672, p=0.006), but not in the Negative-Distance condition (*r_s_*=0.390, p=0.094).

In the Negative-Watch condition, HR was positively associated with negative affect ratings in the FND group (*r_s_*=0.526, p=0.032), but negatively associated with negative affect ratings in HCs (*r_s_*=-0.618, p=0.012), revealing a significant group difference in these coefficients (z=2.92, p=0.004).

HR was negatively correlated with negative affect ratings in the Negative-Distance condition in HCs (*r_s_=*-0.566, p=0.022), but not in FND (*r_s_*=0.321, p=0.143). These coefficients also differed significantly (z=-2.18, p=0.030).

#### Exploratory analyses

No significant relationships were observed between the key experimental outcomes and clinical variables or potential confounds (medication, mental health status).

## Discussion

We aimed to assess the feasibility of an experimental task designed to test the hypotheses that individuals with FND (motor symptoms/seizures) would display elevated autonomic reactivity and increased subjective FNS severity immediately following highly arousing affective stimulation.

### The influence of affective stimulation on subjective FNS

Subjective FNS were significantly elevated immediately after Negative compared to Positive and Neutral blocks. These results concur with two previous studies[19, 20] which showed altered sensorimotor function or subjective FNS in the context of affective processing tasks. However, the experimental design employed by Fiess et al. (2016) did not allow inferences to be made regarding which aspects of the task caused the changes, and Blakemore et al. (2016) measured sensorimotor functioning but not FNS or autonomic arousal.

Our findings provide novel evidence that negative affective events can cause a short-term increase in FNS severity, supporting the proposed role of emotional processing in the generation of FNS[4, 5]. These observations reflect the experiences of many individuals with FND who report that emotionally salient events can trigger or exacerbate their symptoms[6, 8], although these processes may not be applicable in all cases.

### Autonomic reactivity

During the task, the effect of group and the interaction between group x image-type was not significant for SC. Previous findings on task-based SC have been variable across studies in FND samples, with elevated, reduced and comparable SC levels and/or phasic responses reported[4, 18].

There was a significant main effect of group on task-based HR, providing limited support for the hypothesis that the FND group would display enhanced autonomic reactivity. The overall HR deceleration observed in HCs was similar to that observed in other HC samples[29, 33]. In contrast, the FND group displayed overall HR acceleration, corresponding with previous reports[34, 35].

Positive correlations between SC/HR and FNS ratings in the Negative-Watch condition demonstrate a proximate relationship between autonomic arousal during affective stimulation and FNS severity. These findings are compatible with studies showing elevated pre-ictal HR in FS[36, 37], suggesting a role for autonomic arousal as a triggering factor for FNS occurrence/aggravation.

### Intact subjective affect and arousal

The FND group did not differ to HCs in subjective affect and or perceived arousal at any timepoint, despite elevated HR and increased FNS severity ratings following affective stimulation in this FND group. There were also divergent relationships between HR and negative affect ratings in the FND and HC groups during the Negative-Watch condition.

Our results are relevant to models highlighting possible roles for altered interoception and bodily/emotional awareness in the pathophysiology of FND[4, 38, 39], suggesting possible differences in the way that bodily signals of affective arousal might be integrated with negative emotional states in this population.

### The possible influence of voluntary cognitive detachment

In contrast to the Watch condition, subjective FNS severity did not differ between Negative and Neutral blocks in the Distance condition, with FNS ratings elevated to a similar degree in the Neutral-Distance and Negative-Distance conditions. Therefore, during exposure to both affectively neutral and negative events, the experience of cognitive detachment might contribute to the intensity of subjective FNS.

In the Neutral-Distance condition, there were significant positive correlations between SC/HR and FNS ratings, and an inverse relationship between SC and momentary arousal ratings in the FND group. These results indicate that during cognitive detachment in the context of neutral events, greater autonomic arousal was associated with increased FNS severity. Incongruously, this elevated autonomic activation was linked to *reduced* rather than increased *perceived arousal*.

There was a significant correlation between HR and negative affect ratings in HCs in the Negative-Distance condition that was not observed in the FND group. Cognitive detachment might therefore serve to reduce conscious awareness of physiological signals and modulate the experience of negative affect in those with FND[17].

Regarding state dissociation, only dissociative amnesia was elevated in the FND group compared to HCs. These results conflict with elevated trait-depersonalisation scores in this sample, and contrast with previous studies reporting elevations in both detachment and compartmentalisation phenomena in FND[21]. It is possible that some participants found the ‘Distance’ instruction challenging and may have used alternative strategies, such as emotional suppression[40].

### Consistently elevated pain and fatigue

Elevated pain and fatigue ratings in the FND group throughout the experiment support the possible relevance and burden of varied physical symptoms in individuals with FND[24]. Interventions targeting pain, fatigue and other non-FNS somatic symptoms may be critical in FND management.

### Strengths & limitations

Our experimental design allowed us to examine temporal relationships between affective stimulation and momentary FNS severity, alongside objective measures of affective reactivity, offering insights into the possible influence of affective stimulation and autonomic arousal in the pathogenesis of FNS. The experimental model of cognitive detachment is another strength. The influence of potential confounds was eliminated by recruiting HCs who were comparable to the FND group on most characteristics, and we excluded possible relationships between key dependent variables and medication/mental health status.

The study was limited by the small sample size, low statistical power, heterogeneous FND sample, omission of clinical controls and objective FNS assessment. FNS ratings may have been influenced by demand characteristics, although this is unlikely because we did not observe alterations in ratings of other physical symptoms/states, the experimenter provided no information/suggestion about the hypotheses, and the correlations between FNS ratings and autonomic variables indicate that the findings were not merely a result of top-down influences.

### Conclusions

This study provides novel evidence for a possible direct influence of negative affective stimulation on momentary subjective FNS, which was linked to changes in autonomic activation rather than altered subjective affect or perceived arousal. These findings help to unravel the complex influence of affective events on FNS and support models proposing roles for affective/autonomic mechanisms in FND. Interventions aimed at improving awareness, integration and regulation of autonomic signals might offer promise for those with FND.

## Data Availability

Data available on reasonable request.

## Author contributions (CRediT)

Funding acquisition: SP (lead), MH, TC, MAM (supporting). Conceptualisation: SP. Design/methodology: SP (lead), MAM, AATSR, JSW, EW, MJE, LHG, TRN, ASD, TC, MH (supporting). Investigation: SP (lead), BS (supporting). Data curation (processing/preparation): SP (psychophysiology - lead), LSMM (behavioural – lead), EW, ES (behavioural - supporting). Formal data analysis: SP (lead), LSMM (supporting). Supervision: SP (lead), TC, MH, JSW, MJE, LHG, TC, MAM, TRN, AATSR, ASD (supporting). Validation: SP. Project administration: SP. Resources: SP (lead), MH, TC (supporting). Writing: SP (original draft/review and editing), LSMM, TC, MH, MJE, ASD, JSW, LHG, MAM (review and editing). All authors reviewed and approved the final version of the manuscript.

## Acknowledgements

Thank you to our FND Patient and Carer Advisory Panel, all participants, and FND Hope UK and FND Action for supporting the project. Thanks also to Yiqing Sun for contributions to data processing.

For the purposes of open access, the author has applied a Creative Commons Attribution (CC BY) licence to any Accepted Author Manuscript version arising from this submission.

