## Supplementary Materials for "Unravelling the influence of affective stimulation on functional neurological symptoms: A pilot experiment examining potential mechanisms"

### Supplementary Table 1. Eligibility criteria

| | All participants | Functional neurological disorder (FND) | Healthy controls |
|---|---|---|---|
| **Inclusion criteria** | •18-65 years old.<br>•Normal or corrected eyesight.<br>•Fluency in English language. | •A primary diagnosis of FND with either motor symptoms OR seizures.*<br>*Participants with functional seizures were required to experience at least two seizures per month, with premonitory symptoms. | |
| **Exclusion criteria** | •Diagnosis of major cardiovascular disorder (e.g., heart disease).<br>•Diagnosis of major psychiatric disorder (e.g., psychosis, alcohol or substance dependence).<br>•Diagnosis of major neurological disorder (e.g., epilepsy, multiple sclerosis). | •Physical symptoms / disability impairing ability to perform tasks (e.g., severe/constant tremor, bilateral upper limb paralysis, seizure frequency > 10 per day).<br>•Medication that could significantly affect cardiovascular or cognitive functioning (e.g., beta-blockers, high-dose opiates). | •Current diagnosis of any major physical or mental health disorder.<br>•Lifetime diagnosis of functional neurological disorder. |

### Supplementary Table 2. Self-report questionnaire data

| Questionnaire | Description | Scores by group | | Comparison statistics |
|---|---|---|---|---|
| | | FND (Total n=14) | HC (Total n=14) | |
| **Functional Neurological Symptoms Questionnaire** | Assesses the presence, frequency, severity and impact of FNS over the previous week. Scores are calculated for: total number of FNS, average severity (1-7), average impact (1-7), with higher scores indicating more symptoms/greater severity/impact. | See Table 2 | - | - |
| **Patient Health Questionnaire – 15 (Kroenke et al., 2002): M (SD)** | Fifteen items assess the frequency of common somatic symptoms over the previous four weeks. Scores range from 0-30 – higher scores indicate more somatic symptoms. | 13.1 (4.37) | 3.0 (2.32) | t(19.8)=7.68, p<.001, g=2.82 |
| **Patient Health Questionnaire – 9 (Kroenke et al., 2001): Mdn (IQR)** | Nine items measure the frequency of depressive symptoms over the past two weeks. Scores range from 0-27 – higher scores indicate elevated depressive symptoms. | 11.5 (9.0) | 1.5 (3.5) | U=12.5, z=-3.95, p<.001, r=.747 |
| **Generalised Anxiety Disorder – 7 (Spitzer et al., 1999): Mdn (IQR)** | Seven items assess the frequency of generalised anxiety symptoms in the past two weeks. Scores range from 0-21 – higher scores indicate more anxiety. | 7.5 (8.0) | 1.5 (4.5) | U=31.0, z=-3.10, p=.001, r=.586 |
| **Multiscale Dissociation Inventory (Briere, 2002): Mdn (IQR)** | A 30-item measure of the frequency of several forms of psychological dissociation over the preceding month. Raw scores are converted to T-scores (presented here). T-scores range from 0-170 – higher scores indicate greater dissociative symptomology. | **DENG**=60.0 (43.3)<br>**DEPR**=51.5 (48.3)<br>**DERL**=51.5 (30.3)<br>**ECON**=46.0 (10.5)<br>**MEMD**=55.0 (33.8)<br>**IDDIS**=47.0 (0.0) | **DENG**=50.0 (10.0)<br>**DEPR**=47.0 (0.0)<br>**DERL**=46.0 (0.0)<br>**ECON**=46.0 (4.0)<br>**MEMD**=48.5 (8.5)<br>**IDDIS**=47.0 (0.0) | **DENG**: U=57.5, z=-1.88, p=.062, r=.355<br>**DEPR**: U=49.0, z=-2.96, p=.024, r=.559<br>**DERL:** U=56.0, z=-2.33, p=.056, r=.44<br>**ECON:** U=92.5, z=-.296, p=.804, r=.056<br>**MEMD:** U=62.0, z=-1.72, p=.104, r=.325<br>**IDDIS:** U=84.0, z=-1.44, p=.541, r=.272 |
| **Somatoform Dissociation Questionnaire – 20 (Nijenhuis et al., 1996):** | Twenty items examining the extent of various somatoform symptoms in the last year (e.g., sensory disturbances, | 27.5 (9.25) | 20.0 (0.0) | U=11.0, z=-4.22, p<.001, r=.798 |

| | | | | |
|---|---|---|---|---|
| **Mdn (IQR)** | speech/swallowing difficulties, pain). Scores range from 20-100 – higher scores indicate greater somatoform dissociation. | | | |
| **Toronto Alexithymia Scale – 20 (Bagby et al., 1994): M (SD)** | A 20-item measure of difficulties in emotional processing (i.e., identification/description of emotions, external cognitive orientation). Scores range from 20-100 – higher scores indicate greater alexithymia. | 52.5 (11.0) | 41.9 (10.8) | t(26)=2.59, p=.008, g=.949 |
| **Traumatic Experiences Checklist (Nijenhuis et al., 2002): Mdn (IQR)** | A 33-item measure of lifetime traumatic experiences and their impact (e.g., bullying, life threatening illness, childhood abuse and neglect). Due to ethical concerns, we used a 29-item version, omitting the final four items which probe further details of abuse/maltreatment disclosures. Total scores ranged from 0-29 and impact scores for individual events ranged from 1-5. Higher scores signify greater trauma burden and impact. | **Total** =3.5 (5.3) **Impact=**12 (16.3) | **Total=**2.0 (3.5) **Impact=**8.0 (12.0) | **Total:** U=67.0, z=-1.44, p=.164, r=.272 **Impact:** U=67.5, z=-1.41, p=.164, r=.267 |

*Notes.* ECON=emotional constriction; DENG=disengagement; DEPR=depersonalisation; DERL=derealisation; FND=functional neurological disorder; FNS=functional neurological symptoms; IDDIS=identity dissociation; IQR=interquartile range; M=mean; Mdn=median; MEMD=memory disturbance; SD=standard deviation

### Supplementary Table 3. Functional Neurological Symptoms Questionnaire

Please look at the symptoms in the table below and tell us whether you have experienced these functional neurological symptoms in the **past week.** If you mark yes to indicate that the symptom was present in the past week, please complete the additional columns to tell us **how frequent** the symptoms were, **how severe** (intense) they were, and **how much impact** they had on you.

When rating the average **severity** of symptoms, please choose a number from 1 to 7, where **1=Symptom not present; 2=Minimal; 3=Mild; 4=Moderate; 5=Moderately severe; 6=Severe; 7=Very severe**. When rating the **impact** of symptoms, please choose a number from 1 to 7, where **1=No impact at all; 2=Minimal impact; 3=Mild impact; 4=Moderate impact; 5=Moderately severe impact; 6=Severe impact; 7=Very severe impact**.

| **FND Symptom** | **Present?** (circle or bold) | **Frequency** (circle or bold) | **Average severity (1-7)** | **Average impact (1-7)** |
|---|---|---|---|---|
| Weakness | Yes / No | Constant / daily / weekly / less than weekly | | |
| Tremor | Yes / No | Constant / daily / weekly / less than weekly | | |
| Dystonia (muscle spasms / fixed postures) | Yes / No | Constant (1) / daily (2) / weekly (3) / less than weekly (4) | | |
| Walking / mobility difficulties | Yes / No | Constant / daily / weekly / less than weekly | | |
| Myoclonus (muscle jerks) | Yes / No | Constant / daily / weekly / less than weekly | | |
| Seizures* | Yes / No | Number of seizures in the last week: | | |
| Numbness (loss of feeling) | Yes / No | Constant / daily / weekly / less than weekly | | |
| Visual disturbances | Yes / No | Constant / daily / weekly / less than weekly | | |
| Sensitivity to light/sound | Yes / No | Constant / daily / weekly / less than weekly | | |
| Dizziness | Yes / No | Constant / daily / weekly / less than weekly | | |
| Speech / swallowing difficulties | Yes / No | Constant / daily / weekly / less than weekly | | |
| Cognitive difficulties (e.g., brain fog, memory lapses) | Yes / No | Constant / daily / weekly / less than weekly | | |
| Other FND symptoms | Details: | Constant / daily / weekly / less than weekly | | |

Please tell us which FND symptom(s) is most severe and has the most impact on you.

*If you experience FND seizures, do you have warning symptoms? Yes / No

*If you experience warning symptoms before an FND seizure, what is the earliest or most consistent symptom(s) that you experience?

### Supplementary Table 4. Standardised instructions – affective picture task

| | |
|---|---|
| **Instructions 1** | In this task, you will be shown lots of different pictures on the screen, one at a time.<br>There will be 12 blocks of pictures, separated by brief breaks, when the word 'Rest' will be shown on screen.<br>During the breaks, please try to relax and stay focused on the screen.<br>Please press space to continue. |
| **Instructions 2** | Before some of the blocks of pictures, you will see the word 'Watch' on screen for a few seconds.<br>When you see this, please keep your eyes on the screen and just look at the subsequent pictures.<br>Please press space to continue. |
| **Instructions 3** | Before other blocks of pictures, you will see the word 'Distance' on screen.<br>When you see this, please try to minimise your reactions to the pictures by detaching yourself.<br>For example, you could imagine that you are an outside observer, distanced from your personal reactions to the pictures.<br>Please press space to continue. |
| **Instructions 4** | You will see a white fixation cross in the middle of the screen before every picture.<br>You will then see the word 'Watch' or 'Distance' to remind you whether to just look at the picture, or distance yourself from it.<br>You will then see the picture for a few seconds.<br>Some of the pictures may make you feel quite strong reactions; others may not affect you much at all.<br>Please keep your eyes focused on the screen throughout the task.<br>Please press space to continue. |
| **Instructions 5** | After every block of pictures, you will be asked several simple questions about how you feel, right then, in that moment.<br>First, you will be asked about your primary FND symptom, which was specified at enrolment to the study.*<br>You will be asked about different aspects of your current experiences and physical states.<br>You can choose your answers using the number keys on the keyboard.<br>Please try to answer as quickly and accurately as possible.<br>Press space to continue. |
| **Instructions 6** | You will now see some example pictures, with 'Watch' or 'Distance' instructions, so that you can practice the task.<br>Please press space to begin. |
| | **PRACTICE IMAGES x 6** |
| **Instructions 7** | You have now completed the practice items.<br>Please remember to look at the screen and stay as still as possible during this task.<br>Remember, when you see the word 'Watch', just look at the pictures.<br>When you see the word 'Distance', try to minimise your reactions by detaching yourself.<br>Feel free to ask any questions now.[#]<br>Please press space to begin. |

*Sentence omitted from healthy control version

[#]Experimenter answered questions in full and ensured the task was understood fully prior to participants commencing

### Supplementary Table 5. IAPS images

| Image | Description | Category | Mean Arousal | SD Arousal | Mean Valence | SD Valence |
|---|---|---|---|---|---|---|
| 1300 | PitBull | NHA | 6.79 | 1.84 | 3.55 | 1.78 |
| 7380 | RoachPizza | NHA | 5.88 | 2.44 | 2.46 | 1.42 |
| 2811 | Gun | NHA | 6.9 | 2.22 | 2.17 | 1.38 |
| 3064 | Mutilation | NHA | 6.41 | 2.62 | 1.45 | 0.97 |
| 9921 | Fire | NHA | 6.52 | 1.94 | 2.04 | 1.47 |
| 3030 | Mutilation | NHA | 6.76 | 2.1 | 1.91 | 1.56 |
| 9622 | Jet | NHA | 6.26 | 1.98 | 3.1 | 1.9 |
| 2683 | War | NHA | 6.21 | 2.15 | 2.62 | 1.78 |
| 2730 | NativeBoy | NHA | 6.8 | 2.21 | 2.45 | 2.25 |
| 6530 | Attack | NHA | 6.18 | 2.02 | 2.76 | 1.86 |
| 1525 | Attack dog | NHA | 6.51 | 2.25 | 3.09 | 1.72 |
| 2661 | Baby | NHA | 5.76 | 2.13 | 3.9 | 2.49 |
| 1050 | Snake | NHA | 6.87 | 1.68 | 3.46 | 2.15 |
| 3022 | Scream | NHA | 5.88 | 2.08 | 3.7 | 1.91 |
| 3400 | Severed Hand | NHA | 6.91 | 2.22 | 2.35 | 1.9 |
| 6022 | Assault | NHA | 6.09 | 2.47 | 2.14 | 1.55 |
| 9910 | CarAccident | NHA | 6.2 | 2.16 | 2.06 | 1.26 |
| 3102 | BurnVictim | NHA | 6.58 | 2.69 | 1.4 | 1.14 |
| 6415 | DeadTiger | NHA | 6.2 | 2.31 | 2.21 | 1.51 |
| 9620 | Shipwreck | NHA | 6.11 | 2.1 | 2.7 | 1.64 |
| 6312 | Abduction | NHA | 6.37 | 2.3 | 2.48 | 1.52 |
| 1201 | Spider | NHA | 6.36 | 2.11 | 3.55 | 1.88 |
| 2981 | DeerHead | NHA | 5.97 | 2.12 | 2.76 | 1.94 |
| 6212 | Soldier | NHA | 6.01 | 2.44 | 2.19 | 1.49 |
| 9300 | Dirty | NHA | 6 | 2.41 | 2.26 | 1.76 |
| 9500 | Porpoises | NHA | 5.82 | 2.29 | 2.42 | 1.73 |
| 9254 | Assault | NHA | 6.04 | 2.35 | 2.03 | 1.35 |
| 9405 | SlicedHand | NHA | 6.08 | 2.4 | 1.83 | 1.17 |
| 9902 | CarAccident | NHA | 6 | 2.15 | 2.33 | 1.38 |
| 6370 | Attack | NHA | 6.44 | 2.19 | 2.7 | 1.52 |
| 1932 | Shark | NHA | 6.47 | 2.2 | 3.85 | 2.11 |
| 3100 | BurnVictim | NHA | 6.49 | 2.23 | 1.6 | 1.07 |
| 1301 | Dog | NHA | 5.77 | 2.18 | 3.7 | 1.66 |
| 9252 | DeadBody | NHA | 6.64 | 2.33 | 1.98 | 1.59 |
| 3250 | OpenChest | NHA | 6.29 | 1.63 | 3.78 | 1.72 |
| 3150 | Mutilation | NHA | 6.55 | 2.2 | 2.26 | 1.57 |
| 9810 | KKKRally | NHA | 6.62 | 2.26 | 2.09 | 1.78 |

| | | | | | | |
|---|---|---|---|---|---|---|
| 6570 | Suicide | NHA | 6.24 | 2.16 | 2.19 | 1.72 |
| 6315 | BeatenFem | NHA | 6.38 | 2.39 | 2.31 | 1.69 |
| 3063 | Mutilation | NHA | 6.35 | 2.6 | 1.49 | 0.96 |
| 8178 | Cliffdiver | PHA | 6.82 | 2.33 | 6.5 | 2 |
| 7270 | Icecream | PHA | 5.76 | 2.21 | 7.53 | 1.73 |
| 8080 | Sailing | PHA | 6.65 | 2.2 | 7.73 | 1.34 |
| 4689 | EroticCouple | PHA | 6.21 | 1.74 | 6.9 | 1.55 |
| 8341 | WingWalker | PHA | 6.4 | 2.27 | 6.25 | 1.86 |
| 4311 | EroticFemale | PHA | 6.67 | 2.19 | 6.66 | 1.76 |
| 8034 | Skier | PHA | 6.3 | 2.16 | 7.06 | 1.53 |
| 8370 | Rafting | PHA | 6.73 | 2.24 | 7.77 | 1.29 |
| 8186 | SkySurfer | PHA | 6.84 | 2.01 | 7.01 | 1.57 |
| 4677 | EroticCouple | PHA | 6.19 | 2.08 | 6.58 | 1.65 |
| 1650 | Jaguar | PHA | 6.23 | 1.99 | 6.65 | 2.25 |
| 2216 | Children | PHA | 5.83 | 2.2 | 6.41 | 1.9 |
| 8179 | Bungee | PHA | 6.99 | 2.35 | 6.48 | 2.18 |
| 4653 | EroticCouple | PHA | 5.83 | 2.07 | 6.56 | 1.65 |
| 8501 | Money | PHA | 6.44 | 2.29 | 7.91 | 1.66 |
| 8470 | Gymnast | PHA | 6.14 | 2.19 | 7.74 | 1.53 |
| 5621 | Skydivers | PHA | 6.99 | 1.95 | 7.57 | 1.42 |
| 4608 | EroticCouple | PHA | 6.47 | 1.96 | 7.07 | 1.66 |
| 5470 | Astronaut | PHA | 6.02 | 2.26 | 7.35 | 1.62 |
| 8170 | Sailboat | PHA | 6.12 | 2.3 | 7.63 | 1.34 |
| 4695 | EroticCouple | PHA | 6.61 | 1.88 | 6.84 | 1.53 |
| 5626 | Hanglider | PHA | 6.1 | 2.19 | 6.71 | 2.06 |
| 8499 | Rollercoaster | PHA | 6.07 | 2.31 | 7.63 | 1.41 |
| 4664 | EroticCouple | PHA | 6.72 | 2.08 | 6.61 | 2.23 |
| 8190 | Skier | PHA | 6.28 | 2.57 | 8.1 | 1.39 |
| 5450 | Liftoff | PHA | 5.84 | 2.4 | 7.01 | 1.6 |
| 8496 | Waterslide | PHA | 5.79 | 2.26 | 7.58 | 1.63 |
| 7230 | Turkey | PHA | 5.52 | 2.32 | 7.38 | 1.65 |
| 8200 | Waterskier | PHA | 6.35 | 1.98 | 7.54 | 1.37 |
| 4607 | EroticCouple | PHA | 6.34 | 2.16 | 7.03 | 1.84 |
| 8400 | Rafters | PHA | 6.61 | 1.86 | 7.09 | 1.52 |
| 8490 | Rollercoaster | PHA | 6.68 | 1.97 | 7.2 | 2.35 |
| 7502 | Castle | PHA | 5.91 | 2.31 | 7.75 | 1.4 |
| 5629 | Hiker | PHA | 6.55 | 2.11 | 7.03 | 1.55 |
| 4611 | EroticCouple | PHA | 6.04 | 2.11 | 6.62 | 1.82 |
| 8300 | Pilot | PHA | 6.14 | 2.21 | 7.02 | 1.6 |
| 4652 | EroticCouple | PHA | 6.62 | 2.04 | 6.79 | 2.02 |
| 8191 | IceClimber | PHA | 6.19 | 2.17 | 6.07 | 1.73 |

| | | | | | | |
|---|---|---|---|---|---|---|
| 8180 | CliffDivers | PHA | 6.59 | 2.12 | 7.12 | 1.88 |
| 4643 | EroticCouple | PHA | 6.01 | 2 | 6.84 | 1.54 |
| 5531 | Mushroom | Neutral | 3.69 | 2.11 | 5.15 | 1.45 |
| 7009 | Mug | Neutral | 3.01 | 1.97 | 4.93 | 1 |
| 7037 | Trains | Neutral | 3.71 | 2.08 | 4.81 | 1.12 |
| 2396 | Couple | Neutral | 3.34 | 1.83 | 4.91 | 1.05 |
| 2880 | Shadow | Neutral | 2.96 | 1.94 | 5.18 | 1.44 |
| 2516 | Elderly woman | Neutral | 3.5 | 1.88 | 4.9 | 1.43 |
| 2493 | NeutralMale | Neutral | 3.34 | 2.1 | 4.82 | 1.27 |
| 2890 | Twins | Neutral | 2.95 | 1.87 | 4.95 | 1.09 |
| 7207 | Beads | Neutral | 3.57 | 2.25 | 5.15 | 1.46 |
| 7217 | ClothesRack | Neutral | 2.55 | 1.65 | 5 | 0.78 |
| 5510 | Mushroom | Neutral | 2.82 | 2.18 | 5.15 | 1.43 |
| 2102 | NeuMan | Neutral | 3.03 | 1.87 | 5.16 | 0.96 |
| 7059 | KeyRing | Neutral | 2.73 | 1.88 | 4.93 | 0.81 |
| 2514 | Woman | Neutral | 3.5 | 1.81 | 5.19 | 1.09 |
| 7000 | RollingPin | Neutral | 2.42 | 1.79 | 5 | 0.84 |
| 2393 | FactoryWorker | Neutral | 2.93 | 1.88 | 4.87 | 1.06 |
| 7036 | Shipyard | Neutral | 3.32 | 2.04 | 4.88 | 1.08 |
| 2595 | Women | Neutral | 3.71 | 1.88 | 4.88 | 1.24 |
| 7010 | Basket | Neutral | 1.55 | 1.36 | 4.95 | 1.43 |
| 7491 | Building | Neutral | 2.6 | 1.95 | 4.87 | 0.94 |
| 2840 | Chess | Neutral | 2.43 | 1.82 | 4.91 | 1.52 |
| 7235 | Chair | Neutral | 2.68 | 1.9 | 4.85 | 1.13 |
| 7038 | Shoes | Neutral | 3.01 | 1.96 | 4.82 | 1.2 |
| 7055 | LightBulb | Neutral | 3.02 | 1.83 | 4.9 | 0.64 |
| 7160 | Fabric | Neutral | 3.07 | 2.07 | 5.02 | 1.1 |
| 5534 | Mushrooms | Neutral | 3.14 | 2.03 | 4.84 | 1.44 |
| 2385 | Girl | Neutral | 3.64 | 1.81 | 5.2 | 1.32 |
| 7034 | Hammer | Neutral | 3.06 | 1.95 | 4.95 | 0.87 |
| 7950 | Tissue | Neutral | 2.28 | 1.81 | 4.94 | 1.21 |
| 7187 | AbstractArt | Neutral | 2.3 | 1.75 | 5.07 | 1.02 |
| 7179 | Rug | Neutral | 2.88 | 1.97 | 5.06 | 1.05 |
| 9070 | Boy | Neutral | 3.63 | 2.03 | 5.01 | 1.89 |
| 7041 | Baskets | Neutral | 2.6 | 1.78 | 4.99 | 1.12 |
| 2512 | Man | Neutral | 3.46 | 1.75 | 4.86 | 0.84 |
| 5532 | Mushrooms | Neutral | 3.79 | 2.2 | 5.19 | 1.69 |
| 7006 | Bowl | Neutral | 2.33 | 1.67 | 4.88 | 0.99 |
| 7090 | Book | Neutral | 2.61 | 2.03 | 5.19 | 1.46 |
| 6150 | Outlet | Neutral | 3.22 | 2.02 | 5.08 | 1.17 |

| | | | | | | |
|---|---|---|---|---|---|---|
| 2038 | NeuWoman | Neutral | 2.94 | 1.93 | 5.09 | 1.35 |
| 7002 | Towel | Neutral | 3.16 | 2 | 4.97 | 0.97 |

***Notes.*** NHA=negative high arousal; PHA=positive high arousal; SD=standard deviation
